# Quality Improvement (QI) Project: Enhancing Surgical Safety Checklist Utilization at a Rural Primary Hospital

**DOI:** 10.1101/2024.12.06.24318304

**Authors:** Melaku Teshale Gemechu, Anteneh Cheru Adinew, Elias Ermias Ledamo

## Abstract

**Background:** Despite evidence demonstrating various benefits of the World Health Organization (WHO) Surgical Safety Checklist (SSC), its utilization is less frequent in low and middle income countries. This quality improvement project was conducted at a Primary Hospital, a rural facility in Ethiopia, to address suboptimal utilization of the SSC. At baseline, an average SSC utilization rate was 50% and a completion rate was 80% over the prior 6 months.

**Methods:** A single cycle Plan-Do-Study-Act (PDSA) approach was employed to enhance adherence. Key interventions included translating the SSC into Amharic, conducting a two-day training session, introducing pre- and post-operative briefings, and requiring surgeons to remain in the operating room until checklist completion. Bi-weekly audits and regular supervision were conducted to monitor progress. Key performance indicators were SSC utilization and completion rates, while secondary indicators included rate of prophylactic antibiotic administration and surgical site infection. Trend analysis using run charts and Interrupted Time Series (ITS) analysis evaluated changes over time, while staff interviews provided insights into behavioral and attitudinal shifts.

**Results:** The quality improvement project demonstrated a significant and sustained improvement in the utilization and completion rates of the WHO Surgical Safety Checklist (SSC). Baseline measurements in October 2023 showed a utilization rate of 53% and a completion rate of 65%. Following targeted interventions, both metrics steadily improved, reaching 100% by June 2024 and maintaining this level through September 2024.

**Conclusion:** The QI project demonstrated that achieving 100% utilization and completion of the WHO Surgical Safety Checklist (SSC) is feasible even in rural hospitals of low- and middle-income countries (LMICs). Formal training proved crucial for improving adherence, addressing gaps seen with informal approaches, and fostering better team communication and culture. This underscores the value of structured, localized interventions for improving surgical safety practices and encourages broader adoption in similar contexts. To sustain the results, ongoing supportive trainings, monthly supervision, and staff recognition initiatives were initiated.

## Introduction

The World Health Organization (WHO) Surgical Safety Checklist (SSC) has been proven to be effective in reducing surgical morbidity across various settings (1–4). Its adoption in Ethiopian hospitals is a major part of Ethiopian Hospital Reform Implementation Guidelines (EHRIG), and achieving 100 % utilization of the SSC is one of the Ethiopian Ministry of Health’s perioperative quality improvement targets for 2025 (5).

The utilization of SSC is generally high in various countries; however, there is significant diversity across resource contexts and patient groups, even within countries with similar resources. While most facilities are reported to use SSC, it is only performed with reasonable consistency. On average, SSC is used less frequently in countries with a lower Human Development Index and in those where the common or official languages are not among the six WHO official languages. Significant gaps exist in the use of SSC for patients undergoing minor surgeries or urgent surgeries, as well as for obstetric and gynecological procedures (6). Various studies have identified barriers to effective SSC implementation, including the need for changes in the workflow and perception of the checklist and patient safety in general. The factors that hinder or promote the necessary changes are the checklist itself, the implementation process, and the local context, including language, staffing levels, types of surgeries, patient load, and the presence of healthy teamwork (6,7). The absence of positive role models, unenthusiastic team members, hierarchical obstacles, limited knowledge of proper usage, and inadequate implementation procedures have also been reported as barriers to effective SSC adoption (8).

Adherence to the checklist was initially poor in this newly functioning rural facility with an inexperienced operation team. The SSC is often underused or incomplete. This lack of compliance has led to issues such as inconsistent administration of prophylactic antibiotics and frequent cases of lost or mislabeled surgical specimens. Moreover, the inconsistent use of the SSC hindered preparedness for essential surgical instruments and blood products.

Identifying these gaps highlighted the need for a quality improvement (QI) project to enhance adherence to SSC. Implementing a QI initiative would ensure that all team members consistently follow the checklist. Focusing on SSC as part of QI efforts would build a safety culture, improving patient outcomes and efficiency (9,10). This would enhance rate of prophylactic antibiotic administration, reduce specimen handling errors, and ensure preparedness for surgical instruments and blood products.

## Methods

The Hospital ethics committee provided exemption for ethical approval owing to the Quality Improvement nature of this work. Patient consent was also not required for publication. And this work is written in accordance with SQUIRE 2.0 guideline (11).

### Context

The hospital’s surgical unit was staffed by a team that includes one general surgeon, one Obstetrician/Gynecologist, one Integrated Emergency Surgical Officer, two Anesthetists, five Scrub Nurses, and two Cleaners. The hospital provided major surgical service to 226 patients over the last 18 month period.

Despite utilization of the checklist since the start of surgical services, the hospital has not met the national target of 100 % SSC completion over the past 18 months. This gap was a significant challenge in meeting national surgical safety standards. After identifying four areas for improvement in the surgical unit, poor utilization of the SSC was ranked as a priority problem based on a prioritization matrix. Over the past six months, the average utilization of the SSC in the operating room case team was only 50 %, with a completion rate of 80 %. This low adherence rate is likely to lead to increased surgical morbidity. Hence, the project aimed to achieve 100 % utilization and completion of the SSC after three months of intervention; by the end of June 2024.

### Baseline measurement

Initially, data were collected from charts of patients who had undergone surgical procedures over the past six months and these patients were identified from the OR registry. Each chart was reviewed for the utilization and completeness of the WHO SSC, and the findings were recorded using an audit tool. The analysis revealed an average SSC utilization rate of 50 % and a completion rate of 80 %. The results are shown in Figure 1. SSC non-utilization was particularly common in emergency and late-night procedures, with the sign-out section being the most frequently incomplete part of the checklist.

**Figure 1:**
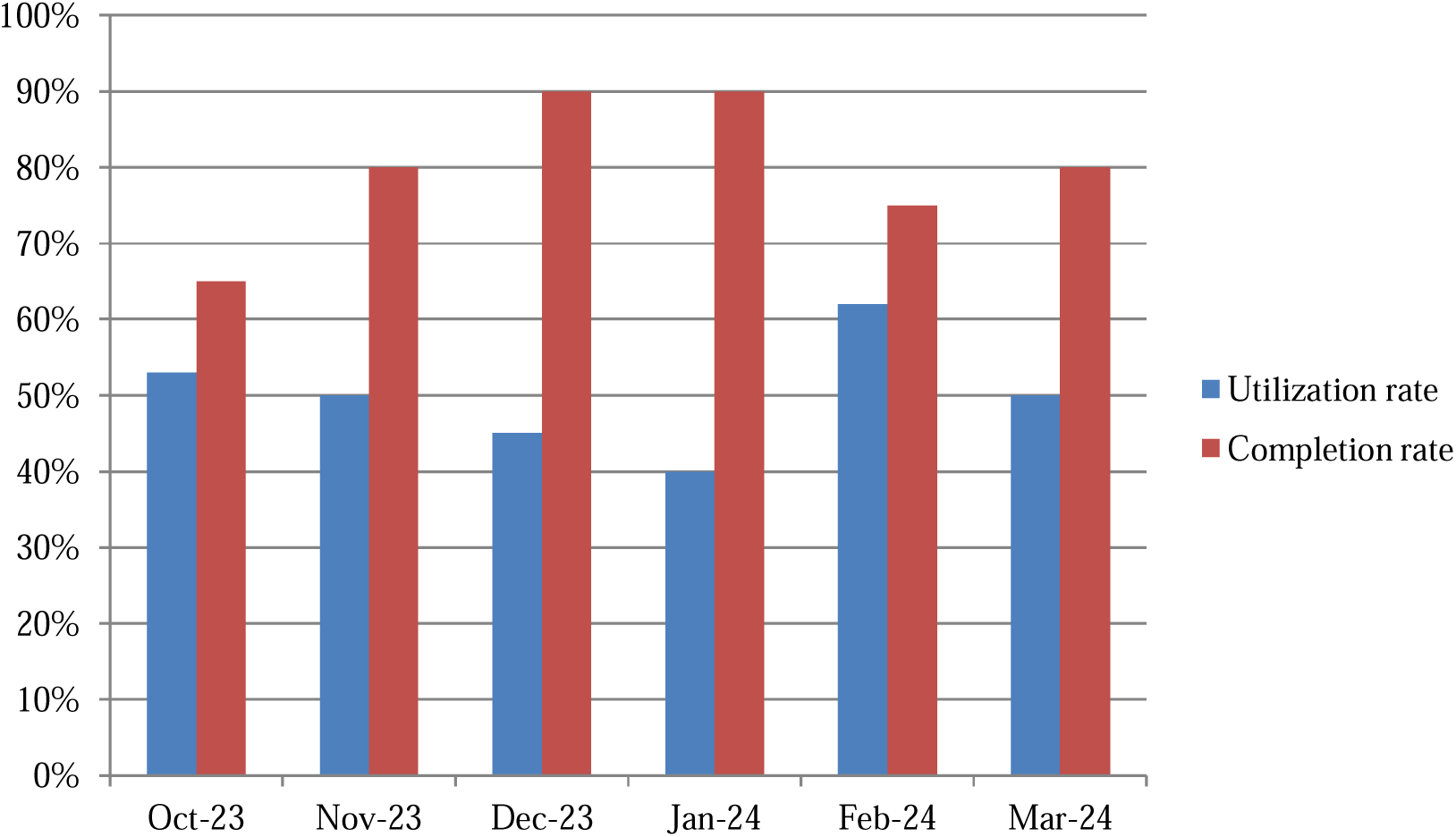
Surgical safety checklist utilization and completeness rate from Oct/2023-Mar/2024.

### Intervention

A driver diagram (figure 2) was used to systematically map out the key factors influencing adherence. We aimed to increase both the SSC utilization rate of 50 % and completion rate of 80 % to 100 % within the following three months. Several primary drivers were identified, including addressing knowledge and attitude gaps, eliminating logistic barriers, increasing leadership commitment, and improving the culture of teamwork. These were further broken down into secondary drivers, such as providing a two days training, commencing a regular preoperative and postoperative briefings, ensuring availability of the SSC hard copy in the OR at all times, and ensuring accountability through monitoring. Change ideas, such as implementing daily reminders for staff, revising the SSC form to make it more user-friendly and translating it into the local language (Amharic) were derived from these drivers. A fishbone diagram (figure 3) that was derived from the driver diagram helped our team to visualize the relationships between these factors. It also enabled us to focus our interventions on areas most likely to influence improvement and guide us through the PDSA cycles.

**Figure 2:**
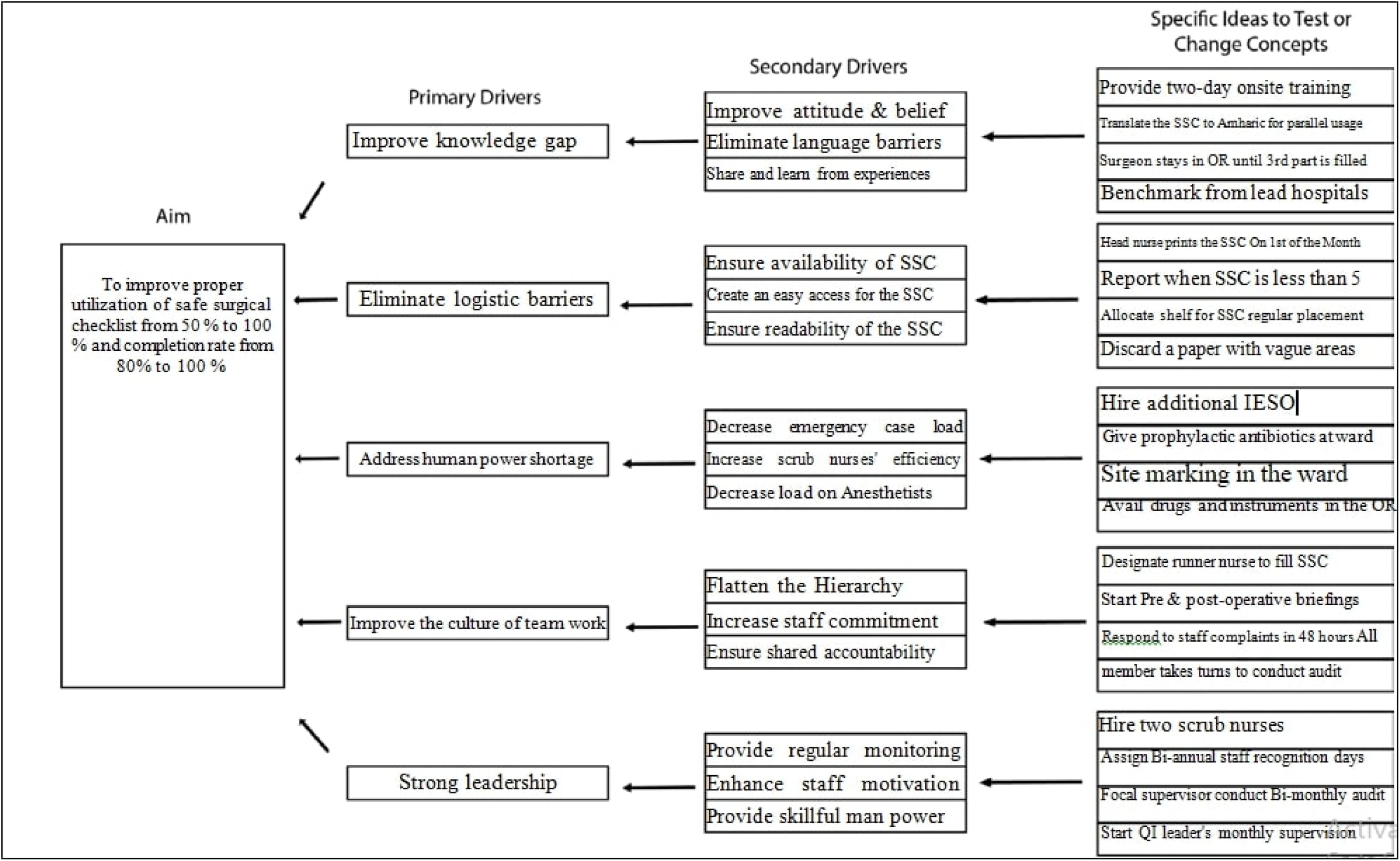
A driver diagram illustrating the primary and secondary drivers, along with the planned change ideas to achieve the target.

**Figure 3:**
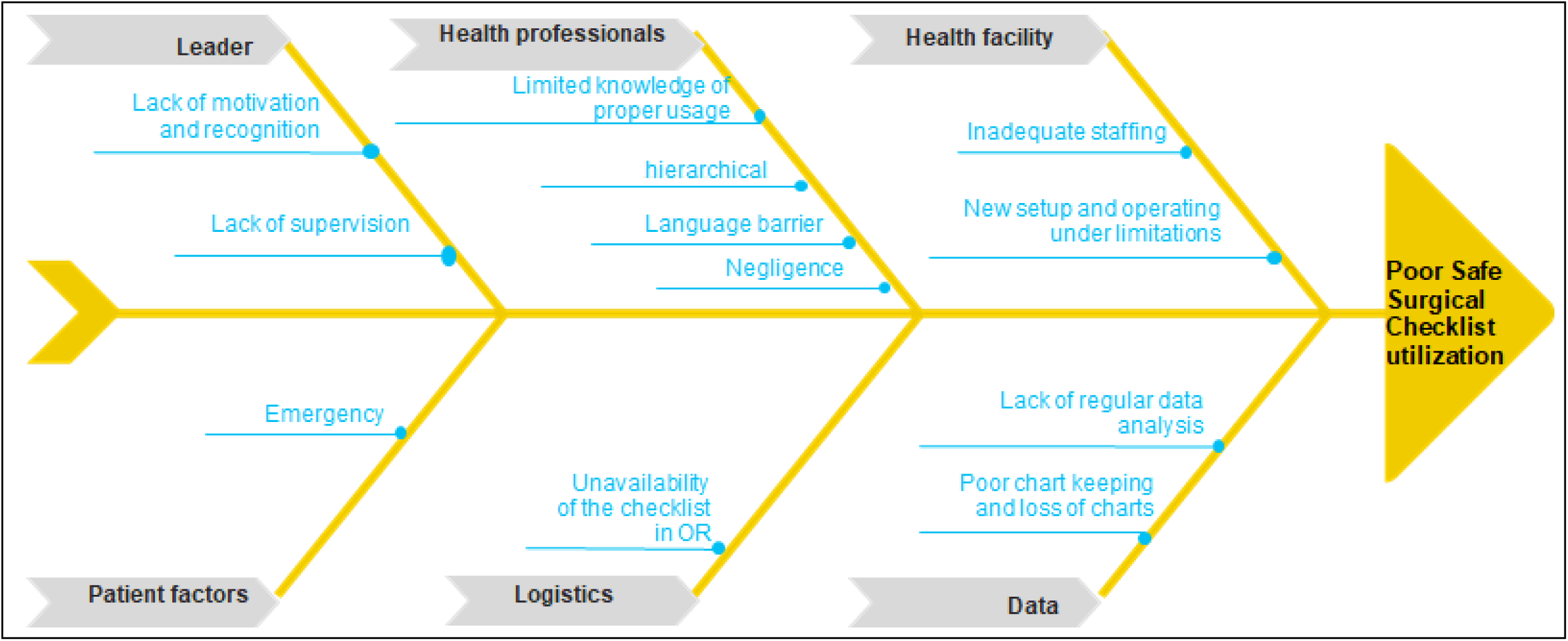
Fishbone diagram depicting factors contributing to poor SSC utilization.

### Strategy

The a single cycle Plan-Do-Study-Act (PDSA) model served as a framework for this project, based on the success of similar past initiatives (9,10). After implementing the intervention strategy, the hospital’s QI director and one operating room (OR) team member audited the compliance and completeness of the SSC every two weeks. Daily monitoring was conducted throughout the three-month study period. Here’s how the PDSA cycle was applied to our QI project with the change ideas listed above:

### Plan

To improve the utilization of the SSC, several targeted interventions will be implemented. The SSC will be translated into Amharic to enhance understanding and facilitate consistent use among staff. Additionally, a two-day onsite training led by a General Surgeon and an Obstetrician/Gynecologist will be conducted to improve staff awareness and skills in using the SSC effectively. To ensure completion, surgeons will be required to remain in the operating room until the third part of the SSC (sign-out section) is completed. The head nurse oversees printing 1.5 times the SSC formats used in the previous month to ensure availability, and ensures the readability of the SSC by discarding incomplete or vague forms. Furthermore, pre- and post-operative briefings will be initiated for every surgical procedure to foster open communication and reduce hierarchical barriers within the surgical team.

To sustain these efforts, shared accountability and regular monitoring are deemed essential. Staff will take turns conducting audits to promote shared responsibility for compliance, and a focal supervisor will perform bi-monthly audits to identify and address gaps. Additionally, the QI director conducts monthly supervision to review SSC adherence and guide improvements. To motivate the team, bi-annual staff recognition days will be introduced to celebrate individuals who excel in SSC compliance.

### Do

Over a three-month period, the team implemented a series of planned interventions to improve the utilization of the SSC. An Amharic-translated version of the SSC was introduced to enhance understanding and facilitate usage among the staff. To build capacity, a two-day training session was provided on February 28 and 29 by a General Surgeon and Obstetrician/Gynecologist, focusing on the importance and proper application of the SSC. Workflows were also adjusted to ensure that surgeons remained in the operating room until SSC was fully completed. Pre- and post-operative briefings were introduced for every surgical case, to foster open communication and address hierarchical barriers.

To monitor the effectiveness of these interventions, bi-weekly audits and regular supervision were conducted. These audits aimed to track compliance, identify gaps, and provide timely feedback. Supervision ensured consistent adherence to the checklist and allowed for the resolution of challenges as they arose. This structured approach helped establish a culture of accountability and collaboration, with the ultimate goal of improving patient safety and surgical outcomes.

### Study

During the implementation period, data were collected through bi-weekly audits to evaluate the effectiveness of the interventions. These audits focused on SSC compliance, utilization, and completeness. A notable improvement in SSC utilization was observed, driven largely by the training sessions and the introduction of pre- and post-operative briefings. Staff interviews confirmed that these changes had an immediate impact, enhancing their understanding of and commitment to the checklist. Adjustments made to workflows also contributed to higher completion rates, particularly during emergency and late-night surgeries.

The audits also revealed an enhanced adherence to the sign-out process, previously identified as the most neglected section of the SSC. This improvement was attributed to the requirement that surgeons remain in the operating room until the checklist was fully completed. Additionally, the pre- and post-operative briefings received a positive feedback from the staff, who noted that these sessions reinforced the importance of teamwork and the SSC.

### Act

Based on the results of the study phase, several actions were taken to sustain the results achieved. Regular training sessions were scheduled every three months to address areas where compliance remained low, ensuring continuous staff development. The Amharic translation of the SSC was extended for an additional three months as a back up to further enhance understanding and completion rates. Monthly supervision by the QI director and bi-annual staff recognition were integrated into the hospital’s formal programs to maintain accountability and motivation. Additionally, the practice of rotating audit responsibilities among all operating room staff continued.

### Study of the intervention

#### Assessing the Impact of the Intervention

1. Key Performance Indicators (KPIs):

- Primary Indicators: Rate of SSC compliance (% of surgeries where the SSC is utilized) and rate of completed SSC (% of fully completed SSC among the utilized checklists).
- Secondary Indicators: The rate of prophylactic antibiotic administration when applicable, surgical site infection (SSI) rates, and rate of lost or unlabeled surgical specimens.
2. Trend analysis was performed using run charts to visualize trends in SSC utilization and completion rates. Interrupted Time Series (ITS) analysis depicting the data collected at multiple time points before and after the intervention will also be used.
3. Interviews with staff were conducted to gather insights into changes in attitudes, behaviors, and perceived outcomes after the interventions.

#### Ethical considerations

- Patient consent for publication: Not required.

## Results

The QI project showed gradual improvement in the utilization and completeness of the WHO SSC over the three months of study period. By June, the project achieved a 100 % rate for both metrics, highlighting the effectiveness of the implemented changes and the team’s commitment to enhancing patient safety in the operating room. Figures 4 and 5 present the results. No missed or unlabeled surgical specimens were reported, and prophylactic antibiotics were administered to all eligible patients during the intervention period. There were instances of unavailable necessary equipment during surgery, primarily due to resource limitations. These issues were addressed by using substitutes or similar items.

**Figure 4:**
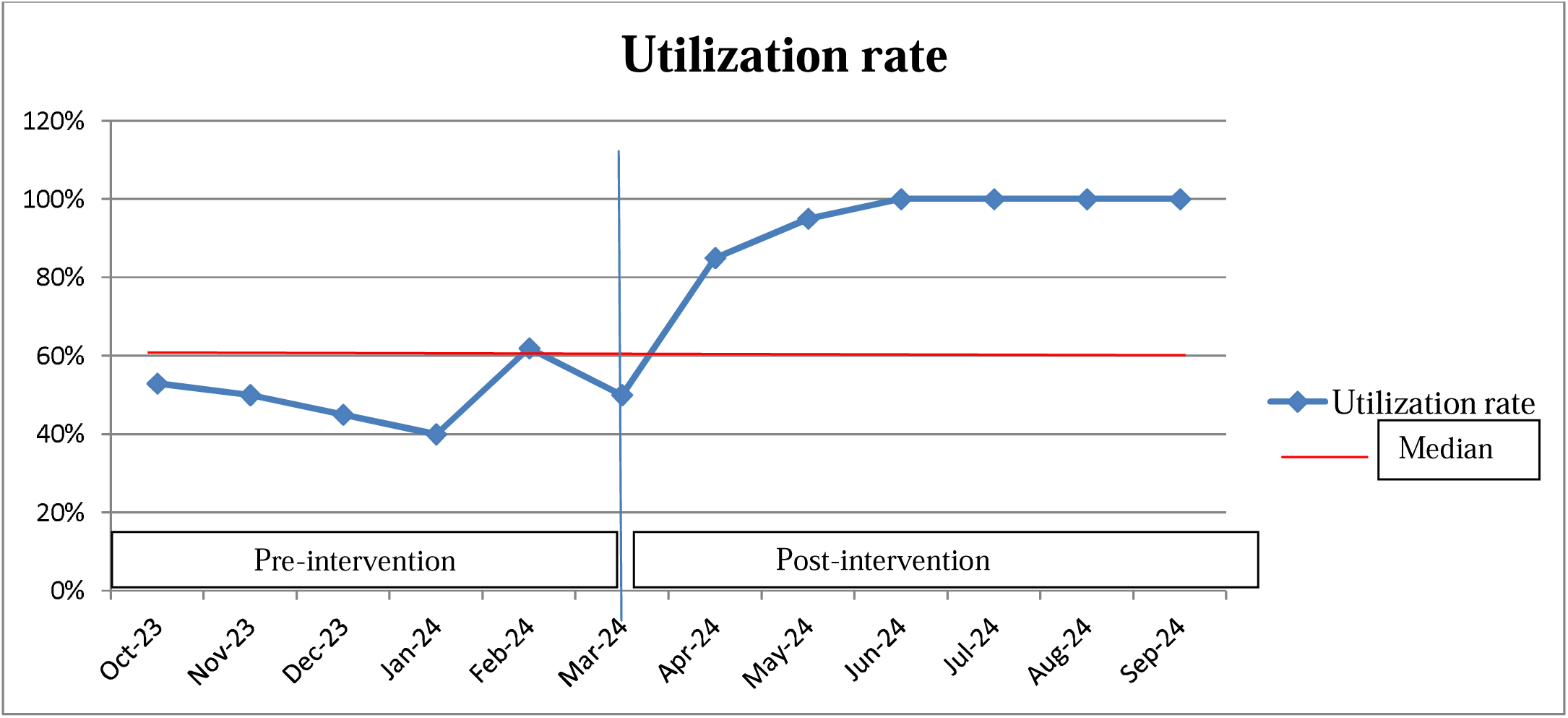
Audit result of utilization rate of the WHO Surgical Safety Checklist from October 2023 – September 2024.

**Figure 5:**
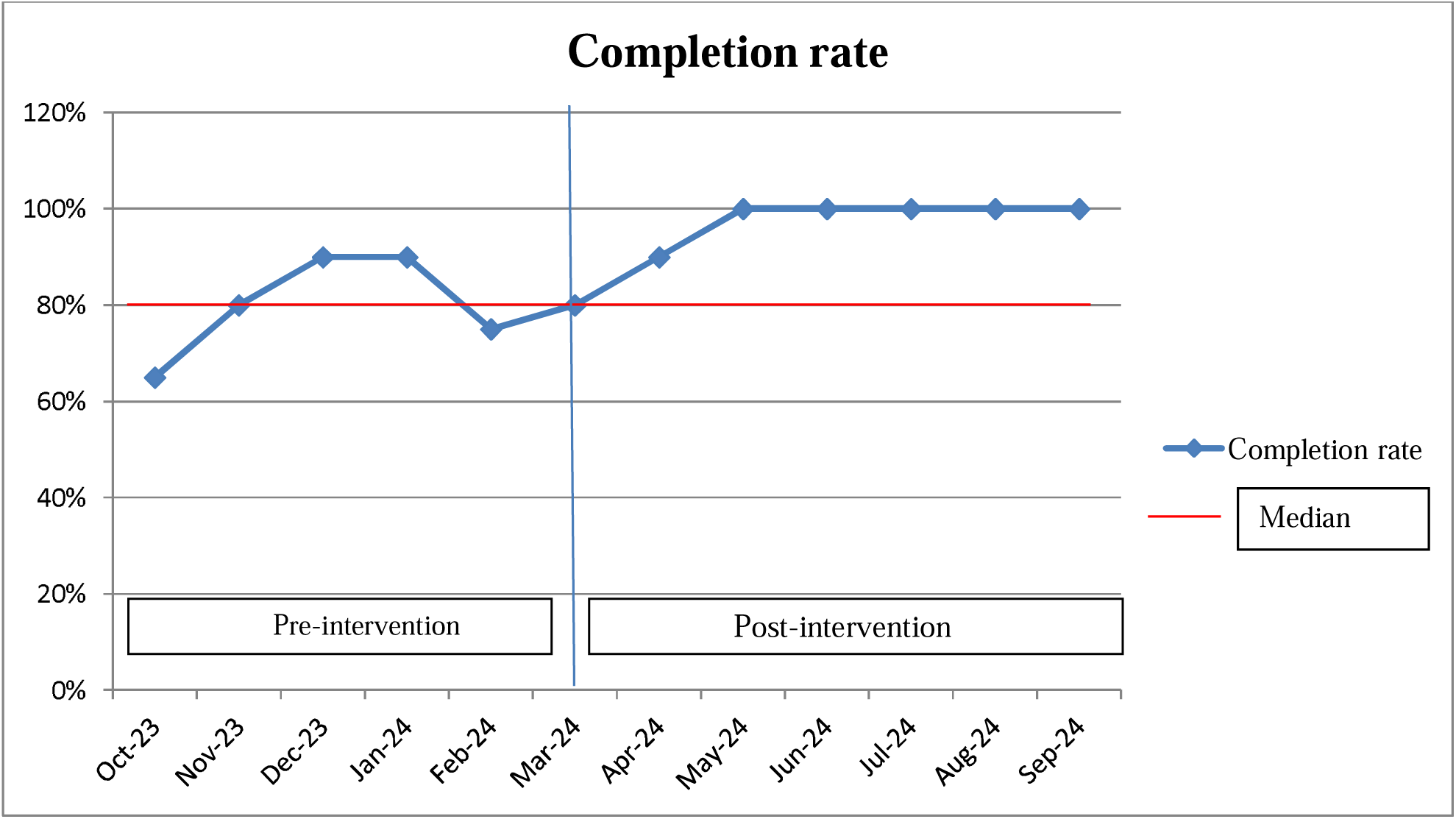
Audit result of completion rate of the WHO Surgical Safety Checklist from October 2023 – September 2024.

## Discussion

The QI project aimed to achieve a 100 % utilization and completion rate of the WHO Surgical Safety Checklist (SSC), based on the assumption of an overall low surgical case burden in the facility and to meet the Ethiopian Ministry of Health’s target. Formal training was essential, as the initial informal approach proved ineffective in improving adherence to the WHO SSC. This finding aligns with various studies that highlight the importance of structured training in enhancing compliance with clinical protocols (8,12–15). Formal training not only increased staff understanding but also reinforced consistent use of the SSC, contributing to the overall success of the QI project.

In low- and middle-income countries, especially in rural hospitals, utilization of the WHO SSC is often low (6,16,17). Language barriers have been identified as a significant contributing factor to this issue, as noted in other studies (6,14). Addressing these barriers by translating the SSC into local languages could facilitate easier implementation and improve adherence.

The “Sign-out” section of the three-part checklist was frequently left incomplete compared to others, primarily due to uncertainty about when it should be filled and because surgeons usually departed the operating room before the checklist was fully completed. Other studies have reported similar results (6,14). While good communication is essential for effective SSC utilization, the reverse was also true in our OR, as team communication significantly improved following the intervention.

Although there is uncertainty regarding the direction of this effect, similar studies supported that proper SSC utilization not only enhances communication but also fosters a stronger culture of teamwork within the surgical environment (14, 18).

Limitations of this QI project include a hawthorn effect and short period of follow up time. The present results reproducibility may be also limited by the influence of local context and institutional culture relating to patient safety.

## Conclusion

This initiative intended to demonstrate that high compliance rates were attainable even in rural hospitals in low- and middle-income countries (LMICs), where utilization rates are typically lower. By successfully achieving 100 % for both utilization and completeness, the project provided a compelling example that adherence to safety protocols can be implemented in these settings. Adopting the checklist in the local context and language, and addressing context-associated barriers played a major role. The results aimed to inspire other rural hospitals to adopt similar practices, ultimately enhancing patient safety and surgical outcomes in regions that are often neglected in terms of healthcare quality.

## Data Availability

All data produced in the present study are available upon reasonable request to the authors.

